# An assessment of coverage and adverse events following country-wide triple-therapy mass drug administration for lymphatic filariasis elimination, Samoa 2018

**DOI:** 10.1101/2020.04.21.20072363

**Authors:** Gabriela A. Willis, Helen Mayfield, Therese Kearns, Take Naseri, Robert Thomsen, Katherine Gass, Sarah Sheridan, Patricia M. Graves, Colleen L. Lau

## Abstract

**Background:** The Global Programme to Eliminate Lymphatic Filariasis is making considerable progress but has experienced challenges in meeting targets in some countries. Recent World Health Organization guidelines have recommended two rounds of triple-drug therapy with ivermectin, diethylcarbamazine (DEC), and albendazole (IDA), in areas where mass drug administration (MDA) results with two drugs (DEC and albendazole) have been suboptimal, as is the case in Samoa. In August 2018, Samoa was the first in the world to implement countrywide triple-drug MDA. This paper aims to describe Samoa’s experience with program coverage and adverse events (AEs) in the first round of triple-drug MDA.

**Methodology/Principal findings:** We assessed MDA awareness, reach, compliance, coverage and AEs from three different data sources: a Supervisor’s Coverage Tool (SCT) in three villages; a large cross-sectional community survey in September/October 2018, 7-11 weeks after the first round of triple-drug MDA; and AE surveillance conducted by the Ministry of Health, Samoa. Participants aged ≥5 years had a fingerprick blood sample tested for circulating filarial antigen using the Alere Filariasis Test Strip. Data were analysed descriptively. In our sample of 4420 people (2.2% of the population), age-adjusted estimates indicated that 89.0% of the eligible population were offered MDA and 80.2% of the total population took MDA. Mild AEs were reported by 13.3% and moderate/severe AEs by 2.9% of participants.

**Conclusions/Significance:** This study following the 2018 triple-drug MDA in Samoa demonstrated a high reported program awareness and reach of 90.8% and 89.0%, respectively. Coverage of 80.2% of the total population showed that MDA was well accepted and well tolerated by the community.

**Author summary:** Lymphatic filariasis is a mosquito transmitted worm disease. A global program underway aims to eliminate lymphatic filariasis as a public health problem by distributing deworming drugs to the whole population once a year for at least five years. In some countries, including Samoa, this strategy has not been sufficient to eliminate transmission. A new drug has been added, and in 2018, Samoa was the first country in the world to apply triple drug mass drug administration using ivermectin, diethylcarbamazine, and albendazole. This study reports on the coverage achieved (percentage of people who reported taking the drugs) and adverse events after taking the drugs. Data were obtained from three different sources. A large community survey of over 4000 people, done 7-11 weeks after the distribution of the first round, found that the program reached and offered MDA to approximately 90% of the whole population, and approximately 80% of the whole population swallowed the drugs. Findings from the community survey on participation in the MDA program were consistent with those from the WHO recommended Supervisor’s Coverage Tool, a smaller survey which was undertaken in three villages by the Samoan Ministry of Health. Data on AEs related to MDA were collected during the community survey, and also through a system set up by the Ministry of Health to enable community members to report any problems related to MDA and receive advice on managing problems. There were relatively few adverse events reported and most of them were mild and of short duration.

## 1 Introduction

Lymphatic filariasis (LF) is a disabling and disfiguring neglected tropical disease caused by infection with three species of filarial worms (*Wuchereria bancrofti, Brugia malayi*, and *B. timori*) [1]. Transmission is by mosquito vectors, which deposit larvae onto the skin when biting humans. The larvae enter the body and migrate to the lymphatic system where they develop into adult worms. Microfilariae (immature larvae), produced by the adult worms, circulate in the blood and infect biting mosquitos, thus enabling ongoing transmission [2]. Chronic manifestations include lymphoedema, typically in the lower limbs, elephantiasis (skin/tissue thickening), and scrotal hydrocoele, which can cause significant disability and social stigma [1, 3]. Laboratory diagnostic tests include detection of microfilariae and circulating filarial antigen to *W. bancrofti* in the blood [4].

In 2014, it was estimated that there were almost 68 million persons living with LF globally, including 36 million microfilaria carriers, 19 million hydrocoele cases, and 17 million lymphoedema cases [5]. LF has been identified by the International Task Force for Disease Eradication as ‘eradicable’ or ‘potentially eradicable’ [6]. The World Health Organization (WHO) launched the Global Programme to Eliminate Lymphatic Filariasis (GPELF) in 2000 with an aim to eliminate LF as a public health problem through community-wide ‘mass drug administration’ (MDA) delivered annually over 4-6 years, which reduces microfilariae prevalence in a population to the point that transmission is considered unsustainable. A second component of the program provides care to people already affected by chronic complications such as lymphoedema and hydrocoele morbidity [7, 8]. Although significant progress has been made, with an estimated 97 million cases of LF being prevented or cured by 2013 [5], the program faces challenges that have slowed progress towards elimination in some countries.

LF is endemic to Samoa, an island country in the South Pacific. Despite delivering 10 rounds of MDA prior to 1999, eight rounds under the Pacific Programme to Eliminate Lymphatic Filariasis (PacELF) (1999-2003, 2006, 2008, and 2011) and two additional rounds in one region of the country (Northwest Upolu 2015 and 2017) [9], a Transmission Assessment Survey (TAS) in 2017 showed evidence of ongoing transmission. According to recently published guidelines, in settings where onchocerciasis is not endemic and where effectiveness of MDA has been suboptimal, as is the case in Samoa, WHO recommends the use of two annual rounds of a triple drug combination (ivermectin, diethylcarbamazine, albendazole [IDA]), a regime shown to be potentially more effective for achieving sustained clearance of microfilariae [10, 11].

In August 2018, Samoa was the first in the world to implement countrywide triple drug MDA. In preparation for MDA, Samoa developed a National Action Plan for Elimination of LF, with the following objectives: i) to stop transmission of LF and prevent new infections; ii) to ensure the provision of basic care for people living with disability due to LF; and iii) to enhance post-MDA surveillance towards validation by 2024. Samoa also established a National LF Control and Elimination Taskforce to oversee preparation, implementation, and monitoring and evaluation of the National Action Plan. The 2018 Samoan campaign aimed to deliver MDA to all eligible individuals through primary and secondary schools, house-to-house visits, workplaces, churches, booths, and central distribution points within communities and ports. Community awareness and advocacy campaigns were conducted through and with schools, workplaces, institutions, churches, and villages. To enhance acceptability of MDA, consultations were conducted to seek engagement and support from multiple stakeholders, including national and local policy-makers, community leaders, religious leaders, school principals, doctors, and ministerial staff [12]

In Samoa, the first round of triple drug MDA was implemented over two weeks in August 2018 by a team of 1600 community drug distributors. A single oral treatment of IDA was given; the number of tablets were calculated based on body weight (ivermectin 150-200μg/kg, diethylcarbamazine [DEC] 6mg/kg, and albendazole 400mg) to determine recommended doses in eight weight categories, and simplified dose charts were used by drug distributors (S1 Table). Directly observed treatment was used whenever possible, and fingernails were marked with indelible ink to indicate participation. MDA was not offered to pregnant women, children aged <2 years, elderly aged >80 years (unless they wished to take the medications), the severely ill, lactating mothers in the first seven days after birth, epileptic children who had experienced a seizure in the previous three weeks, people with heart problems who were experiencing shortness of breath, and people with allergies to any worm medications. Children aged 2-4 years were only offered DEC and albendazole (DA), while children aged ≥5 years and >15 kg were given IDA. Therefore, 2-4 year-olds received two tablets (one DEC and one albendazole), while ≥ 5 year-olds received between three and 17 tablets depending on weight.

MDA coverage in the past has usually been reported as ‘programmatic coverage’, based on summaries of numbers of pills distributed and persons treated from distribution records [13]. There have been few population representative surveys of MDA coverage in the Pacific region. In neighbouring American Samoa, coverage for the 2002 MDA round was estimated to be 54.3% from interviews with 153 participants in a community cluster survey (one person per household, 12 households per village in 20 villages), which was similar to the reported programmatic coverage (49%) [14]. Following the 2004 MDA round in American Samoa, a simple random sample of 1597 persons living in 278 households found a coverage of 81.6%, in comparison to a programmatic coverage estimate of 65% [14]. Achieving high levels of coverage over one or more rounds of MDA is critical to achieving elimination of LF, and taking MDA (also in American Samoa) was significantly associated with reduction in antigen positivity [15].

This paper aims to report on the first round of triple drug MDA in Samoa in 2018, in terms of program awareness, reach, coverage, compliance, and adverse events.

## 2 Methods

### 2.1 Study location

Samoa (previously known as Western Samoa) is an independent country in the South Pacific (latitude 13° 35 South, longitude 172° 20 West) with a population of ∼199,000 [16]. Over 90% of the population live on two main islands: Upolu and Savai’i. Samoa is divided into four administrative regions: Apia Urban Area (AUA), Northwest Upolu (NWU), Rest of Upolu (ROU), and Savaii (SAV). There are ∼338 villages, with average population size of ∼580 (range <20 to 4300) [17].

### 2.2 Data sources

Data on program reach, compliance, coverage, and adverse events were obtained from three separate sources:

A. A population representative survey across Samoa, part of a larger project on Surveillance and Monitoring to Eliminate Lymphatic Filariasis and Scabies from Samoa (SaMELFS Samoa) – conducted 7 to 11 weeks post-MDA, which provided data on reach, compliance, coverage, and adverse events.
B. Supervisor’s Coverage Tool (SCT), conducted in three villages according to WHO guidelines – conducted within two weeks post-MDA, which provided data on reach, compliance, and coverage.
C. Samoa Ministry of Health’s surveillance system on adverse events – during the two weeks of MDA distribution and in the weeks afterwards, which provided data on adverse events.

Each of the data sources is described in detail below.

#### A SaMELFS Samoa 2018

##### Survey design

The Surveillance and Monitoring to Eliminate Lymphatic Filariasis and Scabies from Samoa (SaMELFS Samoa) study was conducted with the primary aims of assessing baseline LF prevalence in Samoa before the first round of triple drug MDA, and to identify ‘hotspots’ of transmission with high antigen (Ag) prevalence (results to be reported in another publication). Due to logistic reasons, the survey was delayed and took place in September/October 2018, 7-11 weeks post-MDA, instead of prior to MDA as intended. Consequently, the SaMELFS 2018 survey was ideally placed to assess the first countrywide use of triple drug MDA. Participants were sampled from 35 primary sampling units (PSUs) located throughout Upolu, Savai’i, and Manono Island (Fig 1). Five PSUs were purposively sampled (three in NWU, one in ROU, and one in SAV) in consultation with the Samoa Ministry of Health, as they were suspected to be transmission ‘hotspots’ based on results of previous surveys. The remaining 30 PSUs were randomly selected using a line list of villages from the 2016 census. Of the 30 randomly selected villages, eight were very small (total population <600) and an adjacent village was added to ensure that target sample size for the PSU was achievable. Therefore, the 35 PSUs included a total of 43 individual villages. The target sample size was 4400, comprising 2000 children aged 5-9 years, 2000 people aged ≥10 years, and 400 children aged <5 years.

**Fig 1.**
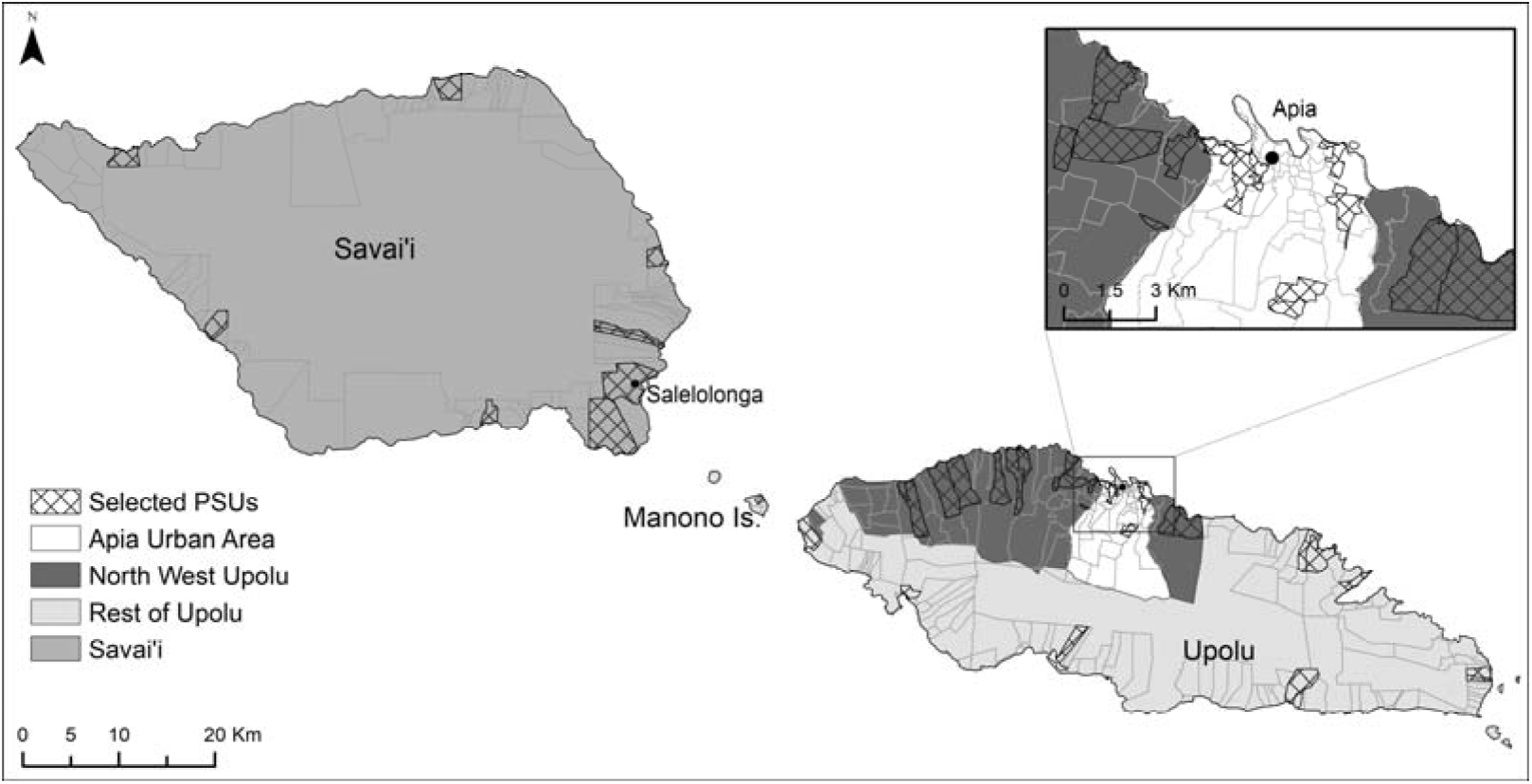
Map of Samoa with administrative region boundaries and selected villages.

##### Data collection

In each PSU, we sampled participants in the selected communities via one of two sampling strategies survey components: i) Household survey (all ages); and ii) Convenience survey (children aged 5-9 years). All participants or parent/guardians (for minors aged <18 years) completed a questionnaire and participants aged ≥5 years had a blood specimen collected and tested for LF antigen and antibodies. These components will be described in further detail below.

###### Household survey

We randomly selected 15 households in each PSU using detailed village aerial maps obtained from Google Maps. All buildings resembling a house were numbered sequentially, and a house was randomly chosen as the starting point for household selection. The remaining 14 houses were selected at equal intervals from this starting point, based on the order in which they were numbered. If a selected household was uninhabited, we replaced it with the nearest inhabited household. If nobody was home at the time of the visit, the house was revisited later in the day and/or when revisiting the village on another day. If household members were still absent at the time of the second attempted visit, we replaced the selected household with the nearest inhabited household.

All household members aged ≥ 2 years were invited to participate in the LF study (children aged <2 years were invited to participate in a linked scabies survey). An individual was considered a ‘household member’ if the house was their primary place of residence, and/or if they slept there the previous night. If eligible household members were not present but were expected to return later in the day, we arranged to revisit the house to include them.

###### Convenience survey

All children aged 5-9 years, who had not been sampled in the household survey, were invited with a parent/guardian to a central place in the village e.g. a school, community hall, church or large *fale* (open house typical of Samoa) for participation in the convenience survey. The number of children enrolled via the convenience survey was dependent on the number of children aged 5-9 years who had already been enrolled via the household survey, with enrolment stopping once the target of 57 had been reached. If insufficient numbers of children attended, we liaised with community leaders to invite more participants.

###### Questionnaires

Interviewers obtained informed consent, enrolled participants and completed questionnaires verbally in Samoan or English, depending on the participant’s preference. Questionnaires were completed by parent/guardians for minors. Data were collected directly into an electronic record on smartphones using Secure Data Kit software (SDK, Altanta, GA). For participants aged ≥5 years we collected data on demographics, information on MDA participation in 2018, reasons for not participating, adverse events, indelible ink marks from MDA participation, and MDA participation in previous rounds prior to 2018. For participants aged 2-4 years, we collected simplified data on demographics and MDA participation in 2018.

###### Specimen collection and testing

For each participant aged ≥ 5 years, we collected up to 400μl of blood by finger prick into a heparin microtainer. Samples were stored in a portable cooler until the team returned to the field laboratory, where they were refrigerated and tested the following day (or on Monday if collected on a Saturday). Blood samples were tested for circulating filarial antigen for *W*.*bancrofti* using the Filariasis Test Strip (FTS; Alere, Scarborough, ME). Positive tests were followed by confirmatory repeat FTS if sufficient blood was available.

##### Data management

We collected enrolment and questionnaire data electronically on smartphones and uploaded regularly to a cloud-based electronic database using SDK, and data were stored on a SQL secure server. Each participant was assigned a unique scannable QR code to link their enrolment/consent form, questionnaire, blood specimen, and laboratory results.

#### B Supervisor’s Coverage Tool

The SCT is one of the current methods recommended by WHO for assessing MDA coverage [18]. This survey requires sampling a single individual from each of 20 randomly selected houses in a supervisory area (such as a village or other administrative unit). Data collected included age, sex, if MDA was offered, if all the pills were swallowed, and where MDA was accessed. The Samoan Ministry of Health had sufficient resources to conduct this survey in three villages in 2018; Faleasiu, Leauva’a, and Nofoali’I. The first two of these villages were also included in the SaMELFS 2018 survey, allowing direct comparisons with the results from the SCT.

#### C Reporting of adverse events to Ministry of Health

Samoa Ministry of Health developed a system for reporting, managing, and investigating adverse events that could potentially be related to MDA. Community drug distributors were provided with training and information to answer common questions from community members. Designated doctors were on call for any adverse events potentially related to MDA and were directly contactable by team leaders if needed. Doctors were also responsible for investigating any severe adverse events and managing risk communication. Media communications were managed with the support of WHO country office in Samoa.

### 2.3 Data analyses

We analysed data using Stata/IC (StataCorp LLC, Texas USA, Version 15.0). A p-value of <0.05 was considered statistically significant. We performed descriptive analyses to explore reported MDA program awareness, reach, coverage, and compliance, as well as reported adverse events. We used Chi-squared tests to compare proportions between population sub-groups and Clopper-Pearson binomial exact methods to estimate 95% confidence intervals (CI). Pearson correlation coefficient was used to measure linear correlation between variables. We used 2011 and 2016 Samoa Bureau of Statistics census data to make demographic comparisons with the general population [19]. Prevalence estimates were standardized for age using the ‘stdize’ option in the ‘proportion’ command in Stata/IC, with ‘stdweights’ as the proportion of the population in each age group (categorized into five-year intervals).

Clustering of coverage was examined using multilevel hierarchical modelling that allowed for correlation of observations by region (n=4), PSU (n=35), and households (n=499) as random effects (Stata command *melogit*). Children from the convenience survey were not included in the models because household-level data were not available. Age and gender were included in the models as fixed effects. Intraclass correlation coefficients (ICCs) with corresponding 95% confidence intervals were obtained from multivariable models.

### 2.4 Ethical and cultural considerations

For the SaMELFS survey, all field activities were carried out in a culturally appropriate and sensitive manner with bilingual local field teams, who received training prior to the study. Verbal approval to conduct the study in the village was sought from community leaders, including the village chief, mayor and/or church leaders. Community leaders disseminated information about the study prior to the visits and assisted with organizing the convenience survey. Prior to enrolment, participants were given verbal information about the study (plus written information if appropriate) in Samoan or English, and we obtained written informed consent from each participant or parent/guardian for minors aged <18 years. For the convenience survey, children were eligible if they had a written consent form from a parent/guardian and were accompanied by a parent or another person (e.g. older sibling or other relative) aged ≥ 15 years. Verbal assent was obtained from minors in addition to parent/guardian written informed consent. Ethical approval was obtained from human research ethics committees at the Samoa Ministry of Health and The Australian National University (protocol 2018/341). The study was conducted in collaboration with the Samoa Department of Health, WHO Samoa country office, Samoa Red Cross, The Task Force for Global Health, and the United States Centers for Disease Control and Prevention. The SCT and adverse events reporting system were part of the Ministry of Health’s programmatic activities for the 2018 round of MDA.

## 3 Results

### 3.1 SaMELFS 2018 study population and antigen prevalence

We recruited a total of 4420 participants from 35 PSUs (43 villages) (∼2.2% of the total population). This included 4211 participants aged ≥2 years who were eligible for MDA: 281 children aged 2-4 years (6.7%), 1941 children aged 5-9 years (46.1%), and 1989 aged 10 years and over (47.2%) (Table 1). Of the 209 participants who were not eligible for MDA, 198 were too young (<2 years of age), seven were pregnant, three were ill, and one did not provide a reason. A total of 2680 participants (63.5%) were sampled via the randomly selected households, and 1542 (36.5%) participants were sampled via the convenience survey. Greater than 90% of households approached agreed to participate. We included a total of 499 households, and an average of 14.3 households per PSU, representing 6.2% of the total estimated 8006 households in sampled villages. Median household size was six people (range 1-20).

**Table 1.**
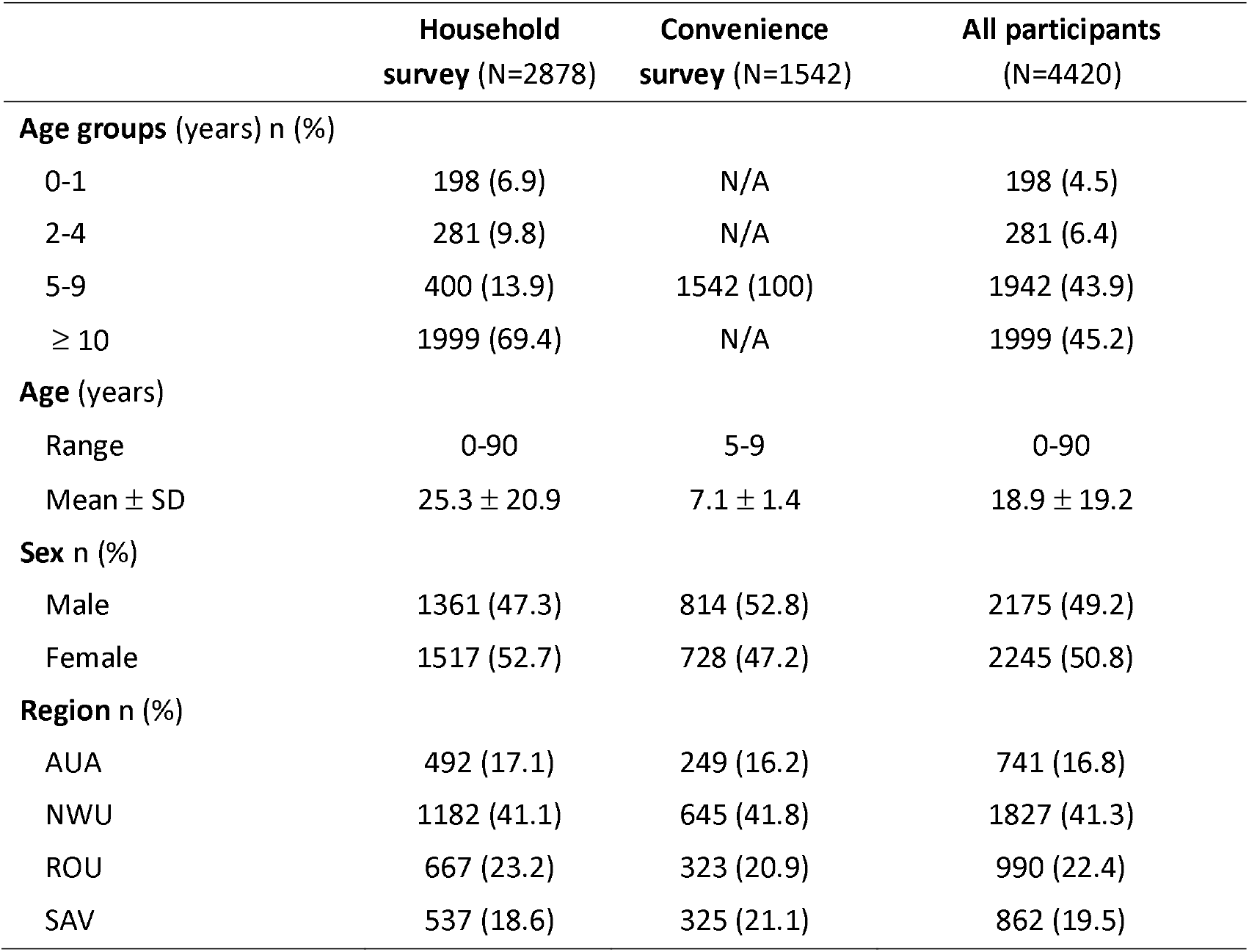
Summary of SaMELFS 2018 study population demographic characteristics.

|  | Household<br>survey (N=2878) | Convenience<br>survey (N=1542) | All participants<br>(N=4420) |
| --- | --- | --- | --- |
| **Age groups** (years) n (%) |  |  |  |
| 0-1 | 198 (6.9) | N/A | 198 (4.5) |
| 2-4 | 281 (9.8) | N/A | 281 (6.4) |
| 5-9 | 400 (13.9) | 1542 (100) | 1942 (43.9) |
| ≥ 10 | 1999 (69.4) | N/A | 1999 (45.2) |
| **Age** (years) |  |  |  |
| Range | 0-90 | 5-9 | 0-90 |
| Mean ± SD | 25.3 ± 20.9 | 7.1 ± 1.4 | 18.9 ± 19.2 |
| **Sex** n (%) |  |  |  |
| Male | 1361 (47.3) | 814 (52.8) | 2175 (49.2) |
| Female | 1517 (52.7) | 728 (47.2) | 2245 (50.8) |
| **Region** n (%) |  |  |  |
| AUA | 492 (17.1) | 249 (16.2) | 741 (16.8) |
| NWU | 1182 (41.1) | 645 (41.8) | 1827 (41.3) |
| ROU | 667 (23.2) | 323 (20.9) | 990 (22.4) |
| SAV | 537 (18.6) | 325 (21.1) | 862 (19.5) |

Age distribution relative to the Samoan population was skewed, with overrepresentation of children aged 5-9 years due to the recruitment strategy and primary study aim of LF surveillance (Fig 2). There was approximately equal sex distribution, with 50.8% of participants being female. Overall, 41.2% were sampled from NWU, with 22.6% from ROU, 16.8% from AUA, and 19.3% from SAV. This was broadly representative of the regional population distribution (35.3% NWU, 23.3% ROU, 19.1% AUA, 22.2% SAV) [17].

**Fig 2.**
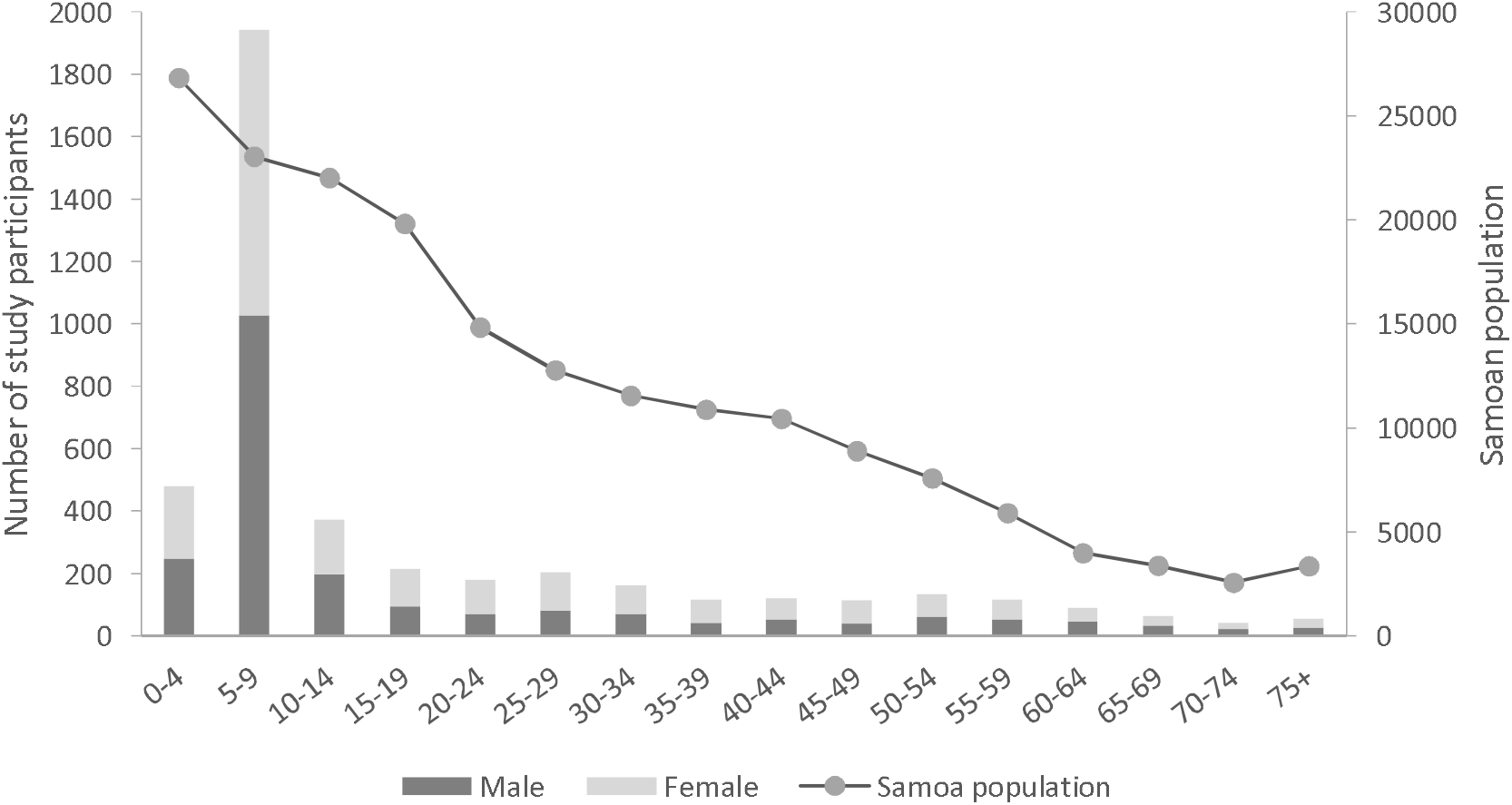
Age distribution of SaMELFS 2018 participants (primary axis) and Samoan population (2011 census) (secondary axis).

Antigen prevalence was 1.5% (95% CI 1.0-2.1%) in participants aged 5-9 years (28 positives out of 1923 valid results), and 4.9% (95% CI 4.0-5.9%) in those aged ≥ 10 years (94 positives out of 1929 valid results). Age-adjusted antigen prevalence was 4.3% (95% CI 3.5-5.2%) for all regions combined, 3.5% (95% CI 2.1-5.7%) for AUA, 6.2% (95% CI 4.9-7.9%) for NWU, 1.8% (95% CI 1.0-3.4%) for ROU, and 3.3% (95% CI 2.0-5.6%) for SAV.

### 3.2 MDA awareness, reach, and coverage

#### 3.2.1 SaMELFS 2018 survey

In participants aged five years and over, 92.4% of participants or their parent/guardian reported being aware of the MDA. The age-adjusted estimate for MDA awareness was 90.8% of those aged ≥ 5 years (or their parent/guardian), highest in ROU (96.4%), followed by AUA (92.0%), SAV (89.6%), and NWU (87.6%). Of those who were eligible for MDA, an age-adjusted estimate of 89.0% were offered MDA (programme reach), and of those who were offered MDA, 99.0% reported taking all the pills (compliance). Age-adjusted coverage was 80.2% of the total population (epidemiological coverage) and 83.9% of the eligible population (programme coverage) (Table 2). MDA coverage was lowest in pre-school children aged 2-4 years (58.4%) and highest in the 10-19 year age group (93.7%) (Fig 3). There was no significant difference in overall coverage (of total population) between males (79.4%, 95% CI 77.4-81.3%) and females (80.8%, 95% CI 79.1-82.4%). Additionally, in children aged 5-9 years, there was no significant difference in coverage rates between randomly selected households (n=400, 93.5%) and the convenience survey (n=1542, 94.3%) (p=0.8).

**Table 2.** MDA awareness, reach, compliance and coverage from SaMELFS 2018.

|  | **Definition** | **Age group used for assessment** | **Sample size** | **Percentage (Unadjusted)** | **Percentage (Age-adjusted)** |
| --- | --- | --- | --- | --- | --- |
| **Awareness** | Proportion of total population that knew about MDA | Participants aged $\geq 5$ years | 3933 | 92.6 | 90.8 |
| **Programme Reach (of eligible population)** | Proportion of eligible population that were offered MDA | Participants aged $\geq 5$ years | 3922 | 91.4 | 89.0 |
| **Programme Reach (of total population)** | Proportion of total population that were offered MDA | Participants aged $\geq 5$ years | 3933 | 91.2 | 88.6 |
| **Compliance** | Proportion of population offered MDA pills who took all pills | Participants aged $\geq 5$ years | 3586 | 99.4 | 99.0 |
| **Programme coverage (coverage of eligible population)** | Proportion of eligible population that swallowed all MDA pills | Participants aged $\geq 2$ years | 4202 | 88.7 | 83.9 |
| **Epidemiological coverage (coverage of total population)** | Proportion of total population that swallowed all MDA pills | All participants | 4411 | 84.5 | 80.2 |

**Fig 3.**
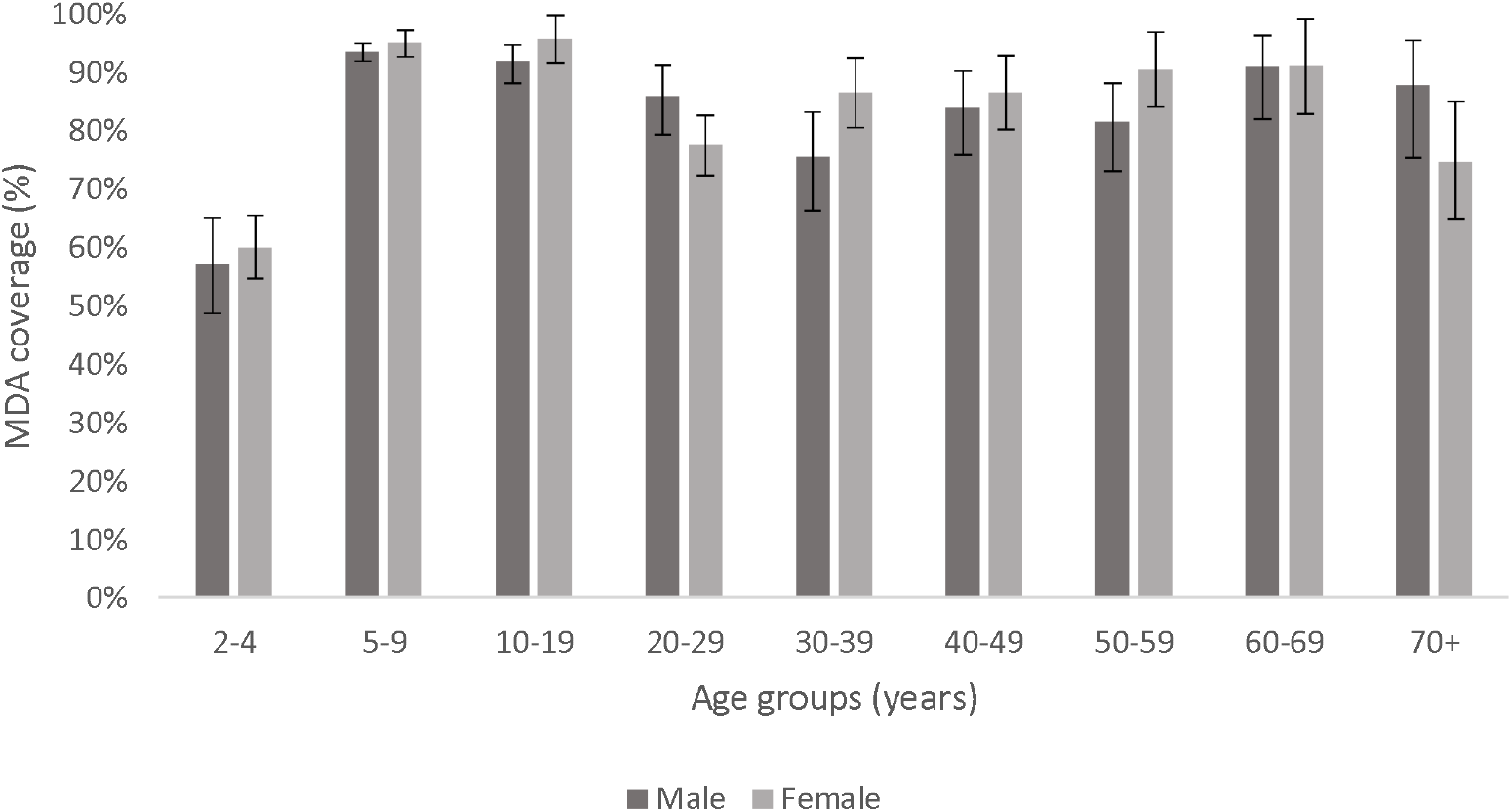
2018 MDA coverage rates (of total population) from SaMELFS 2018, grouped by age groups and sex. Error bars represent 95% confidence intervals.

There was complete concordance in within-household MDA coverage in 60% of households (i.e. household members either all did or all did not take MDA), and in a further 19% of households there was 80 to <100% concordance. The proportion of participants with indelible ink marks gradually declined from 39.1% to 4.1% as the survey progressed, indicating that the ink was fading over time; we were therefore unable to use the presence of ink marks to validate self-report of MDA participation in the SaMELFS survey.

##### Geographical variation in MDA coverage

MDA coverage varied significantly between regions, with the highest age-adjusted coverage rates (of the total population) in ROU (88.1% [95% CI 85.8-90.1%]), followed by SAV (80.3% [95% CI 77.4-82.9%), NWU (78.1% [95% CI 75.9-80.2%]), and AUA (74.6% [95% CI 70.1-77.8%]). At the regional level, there was no significant association between antigen prevalence and coverage. MDA coverage also varied between PSUs, with age-adjusted rates (of total population) ranging from 53.3% to 92.6% (Fig 4). There was no significant difference in age-adjusted coverage rates between randomly (80.5%, 95% CI 79.1-81.8%) and purposively sampled (77.1%, 95% CI 73.4-80.3%) PSUs. At the PSU level, there was a correlation between awareness and coverage (R^2^ 0.68) and reach and coverage (R^2^ 0.86) (Fig 5). Two PSUs in AUA region and one in ROU stood out as having high awareness but relatively low coverage (Fig 5).

**Fig 4.**
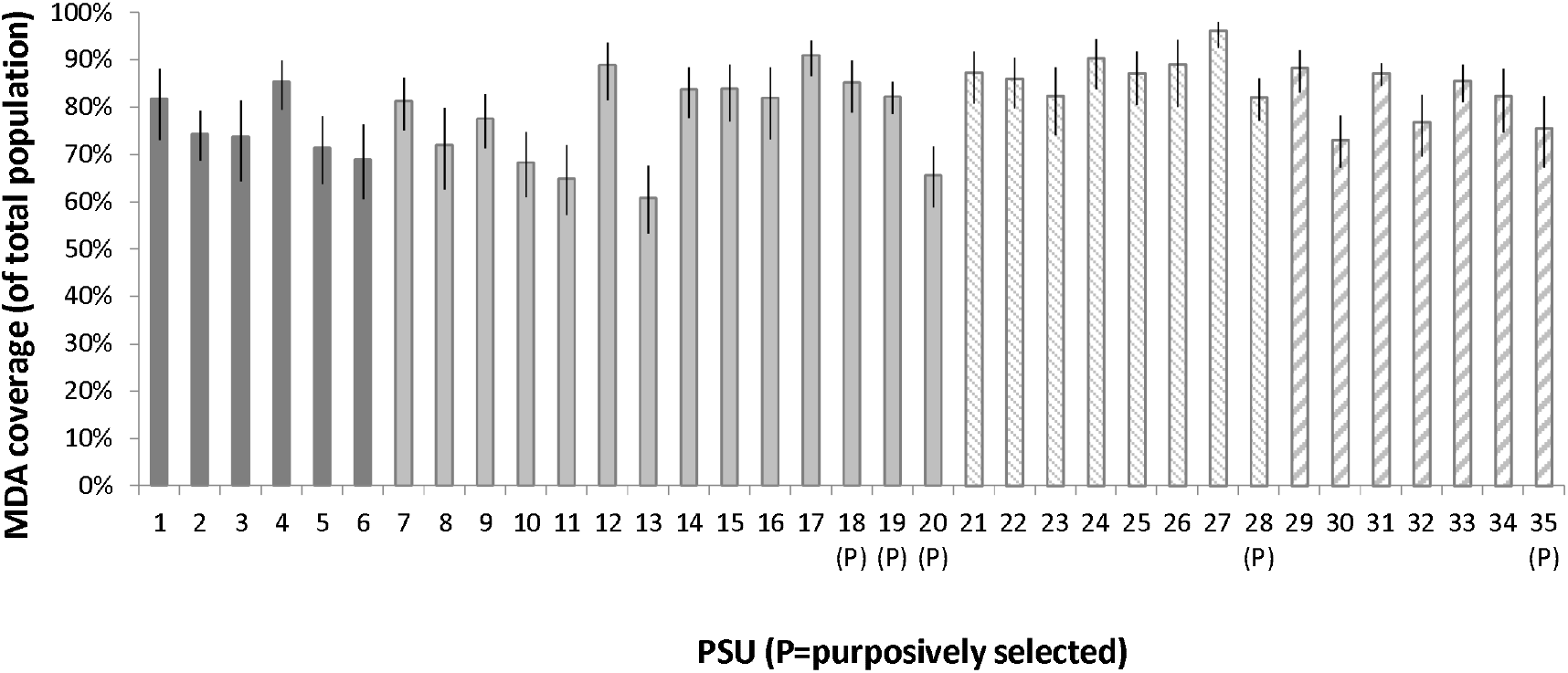
MDA coverage of total population by PSU and Region from SaMELFS 2018.

**Fig 5.**
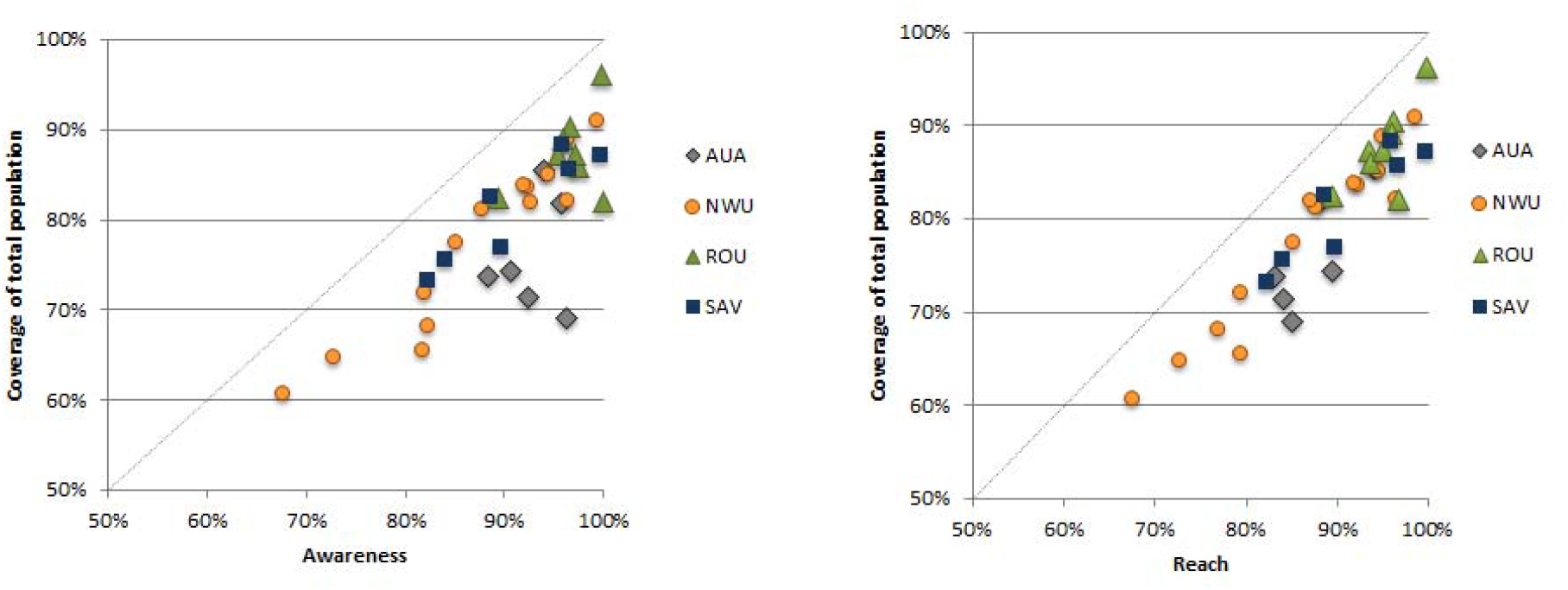
Correlation at each PSU for a) MDA awareness and coverage, and b) MDA reach and coverage, SaMELFS 2018.

##### Intra-cluster correlation

After adjusting for age and gender, ICC at the household level was 0.21 (95% CI 0.14-0.29) for epidemiological coverage and 0.32 (95% CI 0.23-0.42) for programmatic coverage. ICC was highest at the household level, followed by PSU and region, suggesting that coverage was more similar between household members compared to those who lived in the same PSU or region. Figure 6 summarizes the ICCs at region, PSU, and household levels for epidemiological coverage and programme coverage.

**Figure 6.**
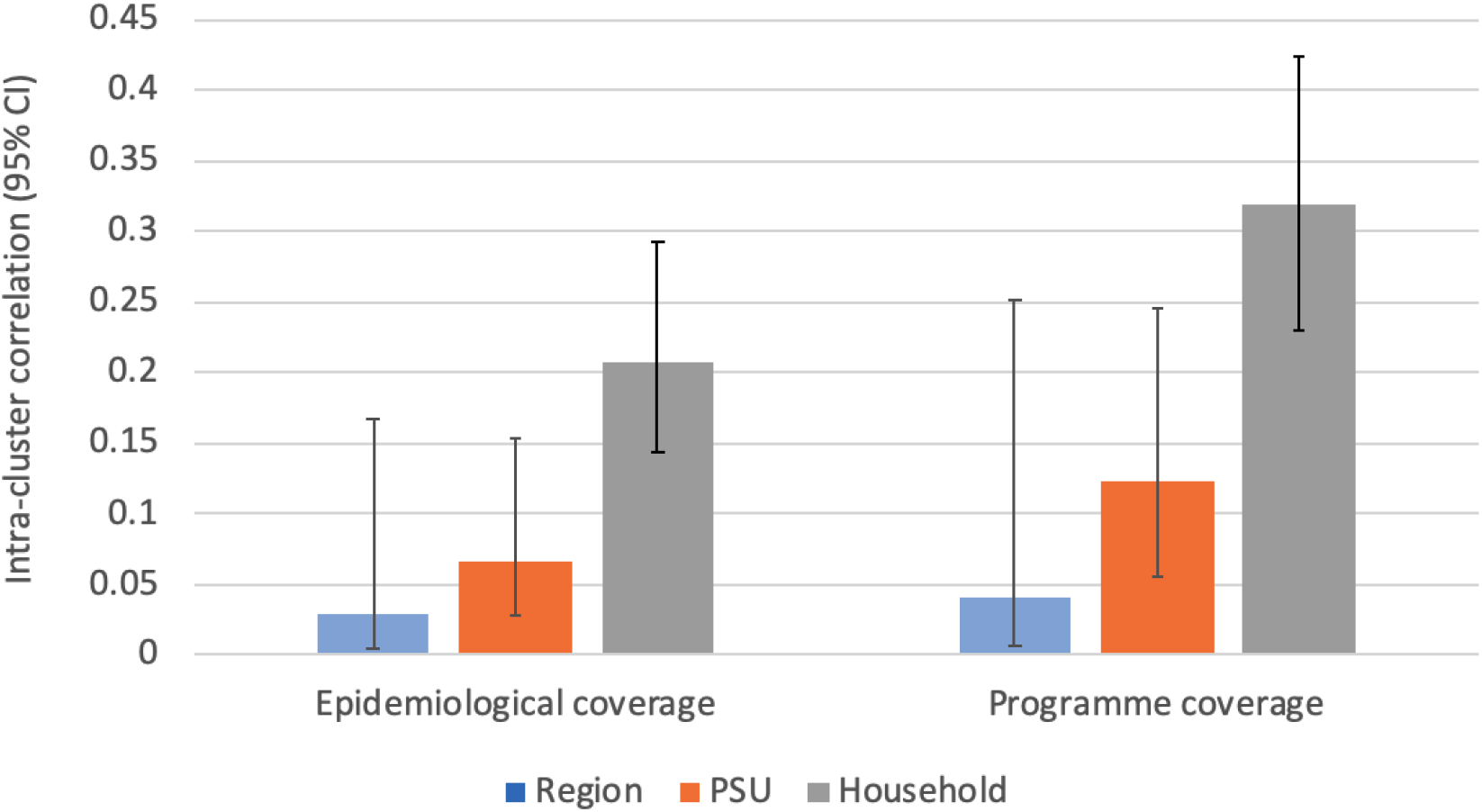
Intra-cluster correlation coefficients for epidemiological coverage and programmatic coverage at region, PSU, and household levels.

##### Reasons for non-participation and non-compliance

Among eligible participants who knew about the MDA (n=3632), 46 participants reported not being offered MDA because the distributors never came, or because they were away, travelling or working. No-one reported that the MDA supply ran out. Among eligible participants who were offered MDA, 18 reported declining. The most common reason given for not wanting to take the tablets was because they were not sick (n=5), being worried about side effects (n=2), didn’t trust the MDA program (n=2), and didn’t like the taste of pills (n=1). Two participants who were offered MDA reported not taking any pills, with reasons given being that there were too many pills (n=1) and no reason given (n=1). Three participants reported taking only some of the pills, with reasons given being that there were too many pills (n=1), they had trouble swallowing all the pills (n=1), and that one pill was lost (n=1).

##### Participation in previous rounds of MDA

In AUA, ROU and SAV regions, where MDA was last conducted in 2008 and 2011, 86.3% of participants aged ≥10 years (i.e. old enough to have previously participated) and who were resident in Samoa at the time, reported participation in MDA in a previous round.

In NWU, where MDA rounds were additionally implemented in 2015 and 2017, among participants who were resident in Samoa at the time, 69.3% aged ≥5 years and 87.4% aged ≥10 years reported participation in previous MDA rounds.

##### Characteristics of people who had never participated in MDA

There were 51 people (2.6% of participants aged 10 years and over) who reported not participating in the 2018 MDA or any previous MDA. Among this cohort, there was equal sex distribution and the highest proportion were in the 10-19 years (33.3%) and 20-29 years (19.6%) age groups. A higher proportion of participants from NWU reported never participating in MDA (3.8%) than from other regions (AUA 0.27%; ROU 1.02%; SAV 1.07%). The most common reason given for not participating in the 2018 MDA was that they didn’t know about it (n=41, 80%). Overall, antigen-prevalence was higher (5.8%) in participants who reported never taking MDA, compared to those who reported taking MDA in 2018 and/or previous rounds (4.9%), but the difference was not statistically significant.

#### 3.2.2 Supervisor’s Coverage Tool

Following WHO guidelines for SCT, 20 people were surveyed in three selected villages within two weeks of MDA, with coverage being 90% or higher in all three villages (Table 3). Reasons for the three people in the SCT survey who reported not being offered MDA and not taking MDA were being away at the time (n=1), being sick (n=1), and parents not being home (n=1). For people who were offered MDA, reported compliance was 100%, but only 84% of those who reported taking MDA had indelible ink on their finger. The percentage with ink marks varied by village from 66.7% to 100%, and reasons given for the absence of ink marks included “scratched it”, “faded”, and “team forgot”.

**Table 3.** Results of SCT in three selected villages.

| Village | Age range | Male n (%) | Offered MDA drugs n (%) | Swallowed MDA drugs n (%) | Where MDA drugs accessed | Finger mark present (% of those who reported taking MDA) |
| --- | --- | --- | --- | --- | --- | --- |
| Faleasiu | 2-70 | 10 (50) | 18 (90) | 18 (90) | 14 Home;<br>2 School;<br>2 Other village | 12 (66.7) |
| Leauva’a | 3-74 | 12 (60) | 19 (95) | 19 (95) | 8 Home;<br>7 School;<br>1 Fixed health facility;<br>3 Other | 19 (100) |
| Nofoali’i | 3-81 | 11 (55) | 20 (100) | 20 (100) | 14 Home;<br>3 School;<br>1 Church;<br>2 Other | 17 (85) |
| **All villages** | **2-81** | **33 (55)** | **57 (95)** | **57 (95)** | **36 Home;**<br>**12 School;**<br>**9 Other** | **48 (84)** |

### 3.3 Reported AEs

#### 3.3.1 SaMELFS 2018 reported AEs

In the SaMELFS survey, of those who participated in MDA and were aged ≥ 5 years, 83.7% reported no AEs, 13.3% reported mild AEs that did not prevent activities of daily living (ADLs), and 2.9% reported moderate or severe AEs that prevented ADLs. AEs were more likely to be reported in those aged 20-29 (22%) and 30-39 (25%) years compared to other age groups (p=0.001), and in females (17.7%) compared to males (15.6%) (p=0.01). Moderate/severe AEs were more common in Ag-positive participants (5.6%) compared to Ag-negative participants (2.8%) who took MDA (Table 4), although this difference was not statistically significant (p=0.149).

**Table 4.** Reported AEs by Ag positivity from SaMELFS 2018.

|  | Ag positivity |  |  | All participants<br>≥5 years* |
| --- | --- | --- | --- | --- |
|  | Ag-positive<br>n<br>(% [95% CI]) | Ag-negative<br>n<br>(% [95% CI]) | Ag unknown<br>n<br>(% [95% CI]) | n<br>(% [95% CI]) |
| **Mild AE** (not preventing ADLs) | 17<br>(15.9 [9.5-24.2]) | 441<br>(13.0 [11.9-14.2]) | 18<br>(25.3 [15.7-37.1]) | 476<br>(13.3 [12.2-14.5]) |
| **Moderate/severe AE** (preventing ADLs) | 6<br>(5.6 [2.1-11.8]) | 96<br>(2.8 [2.3-3.4]) | 1<br>(1.4 [0.0-7.5]) | 103<br>(2.9 [2.4-3.5]) |
| **Any AE** | 23<br>(21.5 [14.1-30.5]) | 537<br>(15.9 [14.6-17.1]) | 19<br>(26.8 [16.9-38.6]) | 579<br>(16.3 [15.1-17.5]) |
| **Total** | **107** | **3385** | **71** | **3563** |

\* Data on adverse events were not collected from participants aged <5 years. ADL= activities of daily living.

#### 3.3.2 AEs from MOH surveillance

A total of 65 persons presented with an adverse event to a public health facility. Reported adverse events were most common in the 2-10 year age group (Table 5) and the most commonly reported symptoms were dizziness, nauseas, lethargy, and a rash (Table 6). No severe adverse events were noted. Four deaths were reported, and immediate investigation determined the causes of death as unrelated to the MDA.

**Table 5.** Age and sex distribution of persons presenting with an AE to a public health facility from MOH surveillance.

| Age group (years) | Male | Female | Total |
| --- | --- | --- | --- |
| 2-10 | 16 | 12 | 28 |
| 11-20 | 8 | 1 | 9 |
| 21-30 | 2 | 2 | 4 |
| 31-40 | 3 | 5 | 8 |
| 41-50 | 2 | 3 | 5 |
| 51-60 | 2 | 1 | 3 |
| 61+ | 5 | 3 | 8 |
| **Total** | **38** | **27** | **65** |

**Table 6.** Presenting symptoms of persons with an AE attending a public health facility from MOH surveillance.

| **Reported symptom (s)** | **Number** | **Percentage (%)** |
| --- | --- | --- |
| Dizziness, lethargy, and nausea | 12 | 18 |
| Dizziness and nausea | 10 | 15 |
| Generalised itchy body rash | 10 | 15 |
| Dizziness, lethargy, nausea and vomiting | 8 | 12 |
| Non-itchy rash | 8 | 12 |
| Malaise and vomiting | 6 | 9 |
| Generalised body pains | 6 | 9 |
| Death during MDA but deemed unrelated to MDA | 4 | 5 |
| Abdominal pain | 1 | 2 |
| Chest pain, shortness of breath, and headache | 1 | 2 |
| **Total** | **65** | **100** |

## Discussion

In 2018, Samoa was the first country to distribute nationwide triple drug MDA, and this paper reports important results on program awareness, reach, coverage, and compliance, as well as adverse reactions related to MDA. This study demonstrates high reported community awareness, reach, and acceptance of MDA in Samoa, likely to be a result of the significant efforts that were put into community awareness and mobilization, advocacy campaigns, and stakeholder engagement. Overall, support of the program was high and mistrust of the program was extremely low, which indicates high community acceptance and a successful education campaign. In other settings, a fear of side effects or poor understanding of LF infection and transmission has resulted in low compliance [7, 20]. Our results demonstrate the importance of both program awareness and reach on coverage, and highlight the importance of community engagement in achieving coverage targets.

In the SaMELFS survey, which included ∼2.2 % of the total population, above the recommended target coverage threshold of ≥65% [11]. We found some geographic variation in awareness and coverage, with NWU having the lowest awareness and AUA the lowest coverage rates. Previous nationwide MDAs in Samoa between 2006 and 2011 reported coverage of 80-90% (Samoa Ministry of Health data). Coverage of MDA targeted to NWU region only in 2015 and 2017 was reported as 70-72% (Samoa Ministry of Health data). The addition of ivermectin and the larger number of tablets with triple drug MDA did not seem to affect compliance, with only three people reporting this as a reason for not participating or not taking all of the tablets. Coverage estimated in the current survey was much higher than previously found in five villages (biased towards high prevalence villages) in Samoa in 2008 after the main series of two drug MDAs by Joseph et al [21]. In that study only 48% of 309 children aged 7-10 years reported taking MDA [21]; however, this surveyed young children in 2008 about MDA participation in 2006 or earlier (when they were less than 8 years old), and results may not be very reliable.

In the current 2018 survey, coverage results from the two villages that were included in both the SaMELFS and SCT surveys were very similar. In Faleasiu, coverage was reported as 95% (114/120) in SaMELFS and 90% (18/20) in the SCT survey. In Leauva’a, coverage was reported as 96.3% (130/135) in SaMELFS and 95% (19/20) in the SCT. Although direct comparison was only possible for two villages, our finding lends weight to the claim that the coverage rates assessed by SCT is comparable to a larger cross-sectional survey methodology, as used in SaMELFS.

For community acceptance of MDA, medications must be well-tolerated and side effects must occur at an acceptably low level. Mild to moderate systemic AEs are common and include fever, headache, dizziness, malaise, myalgia, fatigue and gastrointestinal upset. Localised AEs, thought to arise from the death of adult filarial worms in lymphatic vessels, including subcutaneous or scrotal nodules, spermatic cord swelling, lymphadenitis, or new onset hydrocoele or lymphoedema, occur less frequently [22]. AEs have been observed at higher rates in Mf-positive individuals and are expected to occur at higher rates following MDA in communities with high LF prevalence [22-24]. Given its higher antifilarial activity, it has been postulated that IDA could be associated with higher rates of AEs than DA [23]. Indeed, clinical trials have demonstrated higher AE rates [24, 25], although a recent large multi-center open-label cluster-randomized safety study reported no significant difference in AEs between IDA and DA [23]. We found a self-reported mild AE rate of 13.3% in MDA participants and a moderate/severe AE rate of 2.9%, but no significant difference in rates of AEs in Ag-positive persons. Results from Samoa MOH’s AE surveillance system showed that the most common symptoms reported were dizziness, nauseas, lethargy, and a rash. Although four deaths were reported to the active surveillance system, none were considered related to IDA after thorough investigation by a medical practitioner.

It is difficult to compare our AE occurrence rate from the SaMELFS survey to existing literature, as rates vary significantly depending on method and timing of collection and population characteristics, with passive collection yielding lower rates than active collection. In previous randomized trials of IDA, AEs were reported in 59% [25] and 83% [24] of Mf-positive participants, while Weil et al (2019) found a rate of 12% in their large multi-center safety study with active follow-up [23]. Another recent open-label cohort study of 56 participants reported AEs in 28% of infected (Ag-positive and Mf-positive) and 25% of uninfected (Ag-negative and Mf-negative) individuals, with all reported AEs being mild [26].

A strength of our study was that data were available from multiple sources, which enabled us to provide more detailed information than the standard SCT, and also to cross-validate our results. We found similar results in self-reported coverage rates in the SaMELFS and SCT surveys, and low risk of severe AEs from both the SaMELFS survey and MOH’s AE reporting system. The SaMELFS survey included a large sample size of all age groups across Samoa, and would likely have provided more accurate estimates of coverage compared to the standard SCT, as well as detailed information on differences in coverage between age groups and regions. The SaMELFS survey also assessed program awareness, reach and AEs, which are not generally included in the SCT.

We acknowledge some limitations of our study. The SCT was only undertaken in three villages, and fingernail ink marks were only present in 84% of those who reported taking MDA. In the SaMELFS survey, MDA participation was self-reported 7-11 weeks after the MDA round, and it was not possible to validate self-report by examination of fingernail ink marks as these had largely faded. It is possible that participants did not correctly report MDA participation and/or AEs due to social desirability bias, recall bias, or other reasons. In the SaMELFS survey, children aged 5-9 years old sampled via the convenience surveys may have differed from the general population with higher parental awareness of LF or health literacy, leading to a selection bias. However, the absence of statistically significant differences in reported MDA coverage between children aged 5-9 sampled via the convenience survey and the households, suggests that it would be unlikely for any biases to have affected our results. Although households were randomly sampled, it is possible that households unable to be surveyed due to no one being home when the team visited (and thus replaced with an inhabited household), may also have been more likely to miss the MDA due to working away from home, resulting in an overestimate of the coverage rate. Also, those who were ineligible for MDA (e.g. pregnant, unwell) might have been less likely to agree to participate in the SaMELFS survey, and therefore not included in our estimates of awareness, reach, compliance, and coverage. In the SaMELFS survey, antigen was measured 7-11 weeks after MDA, but results should represent antigen prevalence prior to the MDA [24-26]. A limitation of coverage surveys in general is that results for children are provided by parents or guardians. While we report coverage for children as young as five years, they may not be aware or remember details of the MDA; in young children, our results were of awareness in parents/guardians. Also, parents may not be have been entirely sure about whether children took MDA at school. In the household survey, care was taken to obtain specific answers from each individual, but it was possible that answers might have been influenced by the presence of other household members and their answers.

The success of the 2018 MDA delivery in Samoa was due to a collaborative effort among stakeholders, successful community engagement and mobilization, and a multi-location delivery strategy. The experience in Samoa demonstrates the feasibility and safety of countrywide IDA for the elimination of LF.

## 5 Conclusion

This multi-component study of triple drug MDA implementation and reported adverse events following the 2018 MDA round in Samoa demonstrated a high reported program awareness, reach, coverage, and compliance, and found that the MDA was well accepted and tolerated. Given the need for renewed efforts to eliminate LF using IDA to accelerate GPELF’s progress toward 2030 programmatic goals, our results are encouraging for Samoa’s ongoing MDA program and for other countries that are also planning to implement IDA.

## Data Availability

The data used in the present study are available from the corresponding author upon reasonable request.

## Acknowledgements

We would like to thank all the staff at the Samoa Ministry of Health who supported the many different aspects of the study. We especially Miriama Asoiva who provided valuable advice on local logistics and cultural sensitivities, and assistance with obtaining permissions to conduct village visits. We also thank Fuatai Maiava and Siatua Loau for sharing their knowledge about the LF elimination program in Samoa, and to Fuatai for making it possible for nurses to assist with the household surveys.

We sincerely thank Tautala Maula, the general secretary of the Samoa Red Cross, and her team (especially Babey Suniula, Nixon Mataia, Brenda Koon Wai You, Alesi Mataia, and Shem Lepale) for their enthusiastic and untiring support with fieldwork, village visits, and laboratory work; this survey would not have been possible without their hard work and dedication.

We greatly appreciate all the support and advice provided by Rasul Baghirov and Lepaitai Hansell at the WHO country office in Samoa, and thank them for generously sharing their wisdom. We also thank the Australian volunteers and students who assisted with fieldwork and data management (Brady McPherson, Kelley Meder, Meru Sheel, Benjamin Dickson). We thank the NIH/NIAID Filariasis Research Reagent Resource Center (www.filariasiscenter.org) for supplying positive controls for the Filariasis Test Strips.

## Abbreviations

ADL: activities of daily living
AE(s): adverse event(s)
AUA: Apia Urban Area
Ag: antigen
CI: confidence intervals
DA: diethylcarbamazapine [DEC] and albendazole
DEC: diethylcarbamazine
GPELF: Global Programme to Eliminate Lymphatic Filariasis
FTS: filariasis test strip
IDA: ivermectin, diethylcarbamazine and albendazole
LF: lymphatic filariasis
MDA: mass drug administration
Mf: microfilariae
NWU: Northwest Upolu
PacELF: Pacific Programme to Eliminate Lymphatic Filariasis
PSU: primary sampling unit
ROU: Rest of Upolu
SaMELFS: Samoa Surveillance and Monitoring to Eliminate Lymphatic Filariasis and Scabies from Samoa
SAV: Savai’i
SCT: Supervisor’s Coverage Tool
TAS: Transmission Assessment Survey
WHO: World Health Organization

## Supporting information

S1 Table: Weight-based dosing schedule for triple-drug MDA in Samoa, 2018.

S2 Checklist: STROBE checklist for cross-sectional studies.

